# Evaluation of *in silico tools* for variant classification in missense variants of solid cancer with actionable genetic targets

**DOI:** 10.1101/2024.04.22.24306182

**Authors:** Ilene Hong, Emma Yu, Chiwoo Song, Eugene Kim, Grace Lee, Alice Lee, Young Kwang Chae

## Abstract

Advancement in next-generation sequencing technologies has led to a rise in discovery of variants of uncertain significance, which are not clearly categorized as pathogenic or benign. In silico tools, which have been developed to help classify these variants, exhibit variations in outcome. This study aims to evaluate the performance of 6 widely-used in silico tools in predicting the pathogenicity of drug-actionable gene variants in 9 solid cancers.

We selected drug-actionable genes according to NCCN guidelines on breast, ovarian, colorectal, melanoma of skin, thyroid, bladder, pancreatic, prostate, and biliary cancer. From these genes, we gathered information on 1161 total missense variants (pathogenic = 606, benign = 555). Each variant’s pathogenicity was determined based on assertions from three databases: ClinVar, OncoKB, and My Cancer Genome. We selected variants with one or more concordant databases and excluded variants with conflicting classifications. The performance of the in silico tools (Align-GVGD, CADD, FATHMM, MutationTaster2021, Polyphen-2 (HumDiv), and Polyphen-2 (HumVar)) was evaluated by calculating and comparing their overall accuracy, sensitivity, specificity, and Matthews correlation coefficient (MCC).

Overall, all of the in silico tools demonstrated high sensitivity (0.738-0.927) and moderately-high accuracy (0.555-0.829). Excluding MutationTaster2021, all tools demonstrated low specificity (0.242-0.559) and MCC (0.107-0.413). MutationTaster2021 exhibited the highest performance overall and across solid cancer types. Conversely, Align-GVGD exhibited low performance overall and across cancer types.

Tools demonstrating high sensitivity (CADD: 0.983, MutationTaster2021: 0.927, REVEL: 0.851) could be used to rule out the pathogenic variants. MutationTaster2021, with a comparatively high specificity (0.721), could be considered as an additional test to rule in the pathogenic variants. However, given the varying performance and limitations of the tools according to solid cancer type, clinicians should remain cautious in their usage.

## Introduction

Next-generation sequencing (NGS) is a powerful tool that enables precise and comprehensive genomic analysis in a quick and cost-effective manner [1]. NGS results are frequently used as an indication for certain prognostics and therapeutic options recommended by the National Comprehensive Cancer Network (NCCN) guidelines [1]. The increased utility of NGS in modern medicine has led to the discovery of many variants of uncertain significance (VUS), which are not clearly categorized as pathogenic or benign. Most of these variants are missense substitutions arising from a single nucleotide substitution to the open reading frame [2]. Currently, VUS represent approximately 40% of all identified variants, signifying a large gap in currently-available knowledge [3]. The interpretation based on the differences between wild type and variant amino acids has not been studied sufficiently, leading to errors in clinical judgment [1]. Accordingly, there exist guidelines that provide recommendations on how to interpret sequence variants [4-5]. Most of these guidelines provide five categories based on variant reporting and disease association: benign, likely benign, likely pathogenic, pathogenic, and VUS.

In silico variant classification prediction tools play a pivotal role in clinical settings by offering valuable insights into disease mechanisms, personalized treatment options, and early detection of genetic disorders. However, their variation in prediction algorithms has led to variation in classification of missense variants as pathogenic or benign. The prediction algorithm relies on assumption by two different general approaches: sequence-based and structure-based. The sequence-based approach assumes that differences in amino acid sequence from gene mutation and respective changes in biochemical characteristics of the sequence leads to pathogenicity. Conversely, the structure-based approach uses data derived from 3-D protein structure, including stability and amino acid interactions, to determine pathogenicity [6]. Since each algorithm has a different methodology behind their classifications, the reliability of each tool needs to be tested [7]. There are 6 commonly used in silico tools: Align-GVGD, CADD, FATHMM, MutationTaster2021, PolyPhen-2, and REVEL.

Polyphen-2 inputs eight sequence-based and three structure-based predictive features to its naive Bayes classifier [8]. According to the user’s mutations of interest, either HumDiv- or HumVar-trained Polyphen-2 is selected. The output is the naive Bayes posterior probability a given mutation is damaging, which the tool can then use to categorize the mutation as benign, possibly damaging, or probably damaging [8]. Align-GVGD is a commonly-used tool for BRCA1/2 variant classification. It computes and combines two types of conservation scores, Grantham Variation (GV) and Grantham Deviation (GD), to classify mutations in classes *C* ∈{0,15,25,35,45,55,65}, ranging from least likely to most likely to cause damage [9]. MutationTaster2021, the latest release of MutationTaster, replaces the Naive Bayes classifier of previous versions with Random Forest models for predictions of increased accuracy. The variants are categorized as either deleterious or benign [10]. Functional Analysis through Hidden Markov Models (FATHMM) utilizes a hidden Markov modeling approach that assesses multiple sequence alignments and alignments of conserved protein domain families. This analysis is then used to calculate position-specific amino acid probabilities [11].

While Polyphen-2, Align-GVGD, MutationTaster2021, and FATHMM predict pathogenicity independently, CADD and REVEL are meta-predictors that predict pathogenicity based on a combination of independent variant classification scores. In prior studies, meta-predictors have demonstrated superior performance compared to that of single predictors [14]. Given the clinical relevance of both single and meta-predictive tools, there is a need for greater investigation of their performance and reliability. Our study is the first to extensively evaluate *i*n silico tool performance in nine common solid tumors with actionable genetic targets according to NCCN guidelines.

## Methods

### Cancer Type Selection

We selected solid cancer types with high cancer incidence in the United States and actionable genetic targets according to NCCN guidelines [15-16]. Our selection of 9 common solid cancers included breast, prostate, colorectal, melanoma of skin, bladder/urothelial, pancreatic, thyroid, ovarian, and biliary cancers. Lung cancer was not included because the accuracy of in silico predictors for variants related to NSCLC has been analyzed in a previous study [17].

### Actionable Genetic Target Selection

Actionable genetic targets corresponding to each cancer type were selected according to National Comprehensive Cancer Network (NCCN) guidelines [15]. The actionable gene mutation targets for each selected cancer type are available in Table 1.

**Table 1.**
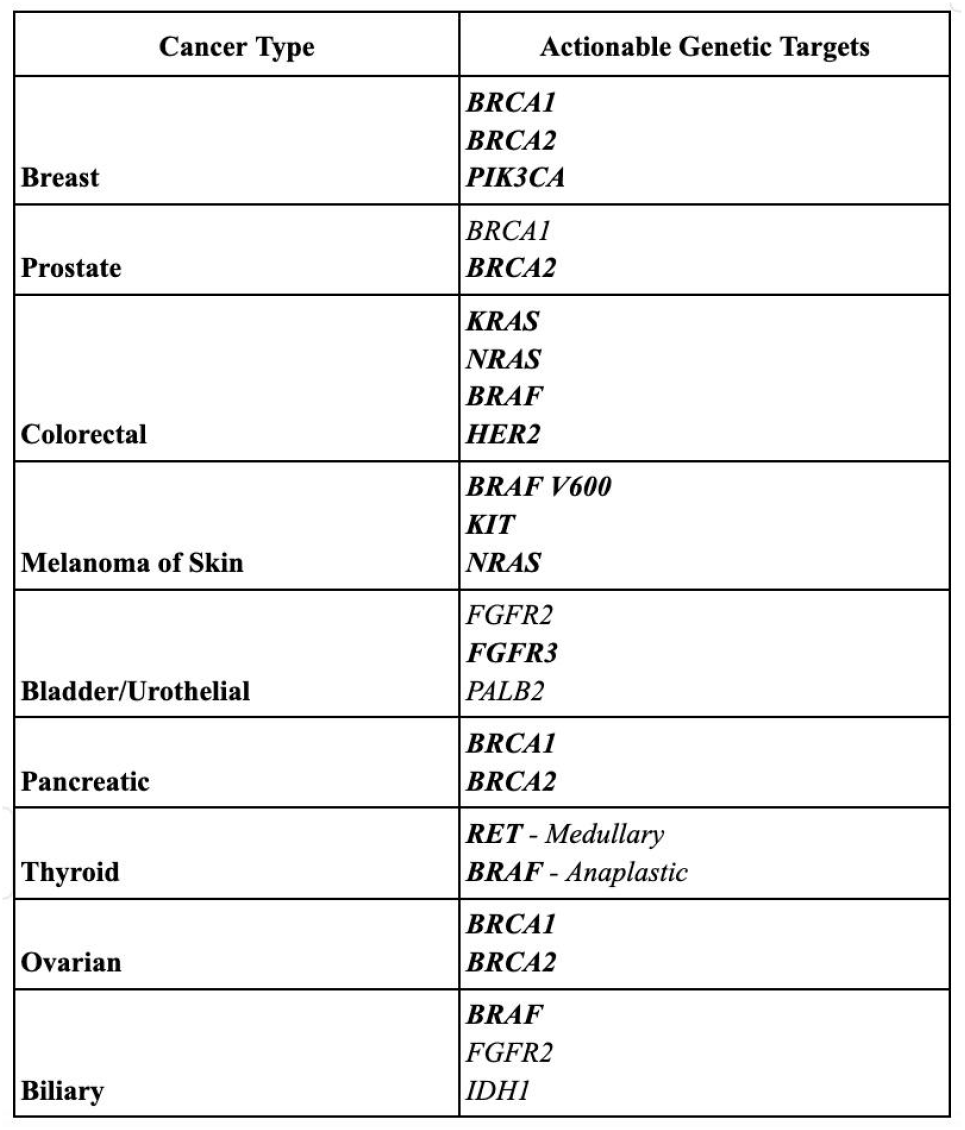
Actionable Genetic Targets According to Solid Cancer Type. Actionable genetic targets of nine cancer types were compiled. Genes in bold lettering were used in analysis.

### Variant Selection

The clinical significance of each variant (benign, likely benign, likely pathogenic, or pathogenic) was determined based on assertions in three databases: ClinVar, OncoKB, and My Cancer Genome. We selected variants with one or more concordant databases and excluded variants with conflicting classifications. To further organize the dataset, we grouped benign and likely benign variants into the “benign” category and pathogenic and likely pathogenic variants into the “pathogenic” category. Variants with conflicting interpretations, uncertain clinical significance, or inconclusive clinical significance were excluded from our analysis. Following the initial dataset compilation, this process of variant collection and classification was repeated to account for potential errors. A final dataset of 1161 missense variants (pathogenic = 606, benign = 555) was curated for analysis.

### In silico Classification Tool Selection

In the present study, we evaluated six in silico tools: Polyphen-2 (consisting of two models: HumDiv, HumVar), Align-GVGD, MutationTaster2021, FATHMM, CADD, and REVEL [8-13]. These commonly-used tools were selected based on their inclusion in the ACMG Standards and Guidelines [4], as well as further literature [17].

### Parameter Setting

We utilized the default thresholds, as established by the tools’ authors, for variant classification. Scores of ≥C35 for Align-GVGD, >15 for CADD,, and >0.50 for REVEL indicated pathogenicity. MutationTaster2021 classifies variants directly as Deleterious, Deleterious (ClinVar), Benign, or Benign (auto).

### Evaluation of Performance

Each of the 652 total missense variants in our dataset was classified using each in silico tool. Correct predictions of pathogenicity were identified as true positive (TP) results, and correct predictions of benign variants were identified as true negative (TN) results. The overall accuracy 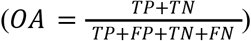, sensitivity 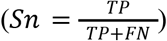, specificity 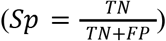, positive predictive value 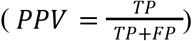, negative predictive value 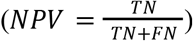, and Matthews correlation coefficient 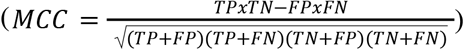 were also calculated for each tool.

Sensitivity (true positive rate) indicated the tools’ ability to identify pathogenic variants, and specificity (true negative rate) indicated the tools’ ability to identify benign variants.

PPV and NPV evaluate the clinical relevance of the tool as it determines the probability of the tool correctly classifying the variants. PPV indicates the probability of the variant being pathogenic after a positive result and NPV indicates the probability of the variant being negative after a negative result.

MCC was used as a balanced performance measure to account for unequal sample size for each prediction tool and proportion of benign to pathogenic variants. MCC values indicate the correlation between predictions and real target values, with a range of -1 (always falsely predicted) to +1 (always perfectly predicted). A value of 0 corresponds to a completely random prediction.

## Results

### Characteristics of selected genes

The pool of genes representing actional genetic targets was narrowed down to BRCA1, BRCA2, and PIK3CA for breast cancer; BRCA1 and BRCA2 for ovarian cancer; KRAS, NRAS, BRAF, and ERBB2 for colorectal cancer; NRAS, KIT, and BRAF V600 for melanoma; RET and BRAF for thyroid cancer; BRCA1 and BRCA2 for pancreatic cancer; FGFR3 for bladder cancer; BRCA2 for prostate cancer; and BRAF for biliary cancer. These genes are presented and bolded in Table 1.

### Characteristics of selected variants

After excluding variants with conflicting classifications (i.e., lacking concordance between databases), the final dataset consisted of 1161 missense variants (pathogenic = 606, benign = 555) across 9 different cancer types. The number of pathogenic and benign variants for each cancer type is available in Table 2.

**Table 2.** Pathogenic and Benign Variant Count by Cancer Type. The number of pathogenic versus benign variants analyzed overall and in each cancer type are displayed.

| Cancer Type | Pathogenic | Benign | Total |
| --- | --- | --- | --- |
| Overall | 606 (52.2%) | 555 (47.8%) | 1161 |
| Breast | 349 (42.4%) | 474 (57.6%) | 823 |
| Ovarian | 259 (36.7%) | 447 (63.3%) | 706 |
| Pancreas | 211 (46.2%) | 246 (53.8%) | 457 |
| Prostate | 210 (46.3%) | 244 (53.7%) | 454 |
| Colorectal | 99 (78.0%) | 28 (22.0%) | 127 |
| Bladder | 63 (63.0%) | 37 (37.0%) | 100 |
| Biliary | 57 (62.6%) | 34 (37.4%) | 91 |
| Melanoma | 67 (77.0%) | 20 (23.0%) | 87 |
| Thyroid | 43 (78.2%) | 12 (21.8%) | 55 |

### Overall Performance

The overall performance of each in silico tool was evaluated based on accuracy, sensitivity, specificity, PPV, NPV, and MCC. Performance scores are highlighted in Supplementary material Table 1. In general, all of the in silico tools achieved high sensitivity (0.738–0.983) and moderately high NPV (0.587–0.947). Specificity was low for all tools (0.242–0.559) with the exception of MutationTaster2021 (0.721). MutationTaster2021 demonstrated the highest overall performance profile with the highest accuracy (0.829), specificity (0.721), PPV (0.784), and MCC (0.666). Superior performance is most evident in accuracy and NPV, where MutationTaster2021 achieved scores >15% higher than other tools. However, CADD outperformed MutationTaster2021 in terms of NPV (0.947) and sensitivity (0.983). Conversely, Align-GVGD demonstrated the lowest overall performance, with the lowest accuracy (0.555), specificity (0.242), PPV (0.548), NPV (0.587), and MCC (0.107).

### Performance of all tools based on solid cancer type

Performance of each tool was evaluated by solid cancer type using the same measures as for overall performance evaluation. Results of this analysis are displayed in Supplementary material Table 1.

In breast cancer variant classification, variants were excluded in each of the tools: Align-GVGD (13/823), CADD (2/823), MutationTaster2021 (9/823), FATHMM (14/823), PolyPhen-2 (HumDiv) (12/823), PolyPhen-2 (HumVar) (12/823), and REVEL (1/823). MutationTaster2021 demonstrated high performance across all performance measurements. CADD achieved the greatest sensitivity (0.986) and NPV (0.972) out of the tools, but had one of the lowest performances for accuracy (0.626) and specificity (0.372). Notably, Align-GVGD achieved a MCC score of 0.114 and the lowest specificity of 0.237.

In colorectal cancer variant interpretation, 2 of 127 variants were excluded in MutationTaster2021. There were no variant exclusions in the other tools. CADD and FATHMM obtained undefined NPV scores, as well as specificities and MCCs of 0. In terms of sensitivity, CADD and FATHMM achieved 100%. MutationTaster2021 achieved scores greater than or equal to 80% across all performance measures, except specificity (0.148) and MCC (0.290). High performance was demonstrated in terms of accuracy, sensitivity, and PPV, with each algorithm achieving scores >75%.

In melanoma variant interpretation, 2 out of 87 variants were excluded when observed with MutationTaster2021. CADD and FATHMM obtained undefined NPV scores, as well as specificities and MCCs of 0. CADD, FATHMM, and MutationTaster2021 achieved sensitivity scores of 100%. Each algorithm achieved a score >75% for accuracy, sensitivity, and PPV.

In ovarian cancer variant classification, variants were excluded for Align-GVGD (13/706), CADD (2/706), MutationTaster2021 (7/706), FATHMM (14/706), PolyPhen-2 (HumDiv) (13/706), PolyPhen-2 (HumVar) (13/706), and REVEL (2/706). MutationTaster2021 demonstrated the greatest overall performance and achieved the highest scores across the performance measures, excluding sensitivity (0.907) and NPV (0.939). CADD achieved the greatest sensitivity (0.988) and NPV (0.982), but did not display high performance in other performance measures. Align-GVGD achieved the lowest scores for MCC (0.121) and specificity (0.233).

In pancreatic cancer variant classification, variants were excluded for Align-GVGD (1/457), FATHMM (2/457), and MutationTaster2021 (3/457). MutationTaster2021 demonstrated high performance across all performance measures but was outperformed by CADD in terms of sensitivity (0.895 vs 0.976) and NPV (0.892 vs 0.932). Align-GVGD resulted in a low specificity and MCC, once again, with scores of 0.220 and 0.121, respectively.

In prostate cancer variant classification, variants were excluded for Align-GVGD (1/454), FATHMM (2/454), and MutationTaster2021 (3/454). Again, MutationTaster2021 demonstrated the highest performance across all performance measurement, except sensitivity and NPV. While MutationTaster2021 achieved sensitivity and NPV scores of 0.895 and 0.891, CADD achieved higher scores of 0.976 and 0.922. Align-GVGD and FATHMM achieved the lowest MCC scores of 0.125 and 0.143, respectively.

In biliary cancer variant classification, variants were excluded for Align-GVGD (8/91). MutationTaster2021 demonstrated high performance across all performance measurements, excluding sensitivity (0.877 vs 0.982) and NPV (0.500 vs 0.667), where it was outperformed by CADD. Specificity scores for all tools is low, with a narrow range of 0.059 to 0.265. All tools also had low MCC scores, ranging from 0.021 to 0.112.

No variants in thyroid cancer variant interpretation were excluded. MutationTaster2021 and REVEL performed highly in all performance measures. Each algorithm demonstrated high sensitivity and PPV, with scores >75%. Notably, FATHMM obtained an undefined NPV score, as well as specificity and MCC of 0. However, it achieved a sensitivity of 100%.

## Discussion

This study evaluates in silico tool performance for predicting pathogenicity of missense variants of multiple solid tumors with actionable genetic targets. Our findings indicate MutationTaster2021, an individual predictor, as the highest performer in predicting pathogenicity. This deviates from prior studies indicating superior performance of meta-predictors, such as REVEL and CADD, over individual predictors [18-19].

CADD demonstrated the highest or second highest sensitivity in overall analyses and across each cancer type. This highlights its ability to effectively detect a higher proportion of deleterious variants. However, CADD also exhibited consistently low specificity (<0.4) across each cancer type. Its overall specificity was calculated as 0.319, which is 55.7% lower than 0.721, the overall specificity of MutationTaster. This suggests that CADD may be useful in detecting truly deleterious variants but at the cost of a higher probability of false positives. It is worth noting that CADD and FATHMM exhibited specificities and MCC of 0.00 and undefined NPV in colorectal, melanoma, and thyroid cancer analyses. This can be attributed to the absence of true negative and false negative classes in our dataset, and thus our analyses of CADD and FATHMM are inconclusive for these cancer types.

REVEL demonstrated on par or high performance across each performance metric compared to the other tools. Since it does not demonstrate notably higher performance, we suggest that is not the optimal tool. Like CADD, REVEL resulted in high sensitivity and low to moderately-low specificity in each cancer type. Previous studies have shown REVEL to outperform other in silico tools, including CADD, but our results indicate that REVEL may not be as accurate in predicting pathogenicity of variants corresponding to certain solid cancer types in this study. However, since REVEL did not result in an undefined NPV or specificity of zero in colorectal cancer or melanoma, REVEL may be the more useful meta-predictor tool for classifying variants of uncertain significance in these cancer types.

Align-GVGD’s worst overall performance in this study aligns with a previous study investigating in silico tools for variant classification in clinically actionable NSCLC variants [17]. Moreover, Align-GVGD resulted in an MCC greater than zero and less than 0.2 in all cancer types, which suggests that the model’s performance is better than random chance but is still significantly limited in its predictive ability. This further supports its inferior performance and its inability to handle imbalanced class distributions.

In our study, MutationTaster2021, launched on 07/02/2021, stands as the newest tool [10], while Align-GVGD, last updated on 09/18/2014, stands as one of the oldest tools [9]. The contrasting results between the latest and oldest tools in our investigation underscores the rapid advancements in precision genomics and emphasizes the importance of tools staying current to accommodate the expanding knowledge of pathogenic variants. Additionally, our study utilizes FATHMM-MKL, a model launched in 2013 [11]. The updated version, FATHMM-XF, launched in 2018 may be worth investigating in future studies [20].

Clinical relevance of in silico classification tools is demonstrated by the dependency of biotechnology companies on these tools in assessing patient samples. Our study reveals a number of issues that may potentially limit usage of these tools in clinical settings. First, a large degree of variability was observed in tool performance among missense variants of different solid cancer types with actionable genetic targets. False negative and/or false positive predictions pose risk for subpar, and thus ineffective, treatment for patients, which may result in exacerbation of medical conditions and elevation of healthcare expenses. Additionally, our research exclusively focuses on missense variants because the majority of variants pertinent to our selected cancer types are missense substitutions arising from single nucleotide substitutions in the open reading frame [3]. However, deducing the functional effects of missense variants is more complex compared to frameshift or nonsense variants [21]. Furthermore, the relatively low sample sizes for colorectal, bladder, biliary, melanoma, and thyroid cancers and the relatively high class imbalance (Table 2) should be taken into consideration when interpreting results for these specific cancer types. Despite its limitations, our findings underscore the nuanced nature of in silico tool performance in different cancer types.

Although in silico tools capable of analyzing mutations beyond missense mutations exist, such as DANN [22] and DeepSEA [23], a tool that can assign pathogenicity to mutations across mutation classes remains necessary. Our study also highlights the potential value of a single, comprehensive in silico variant classification tool that displays high performance in predicting pathogenicity of mutations across solid cancer types. Until these needs are met, clinicians should exercise caution with in silico tools when evaluating actionable gene targets to aid their clinical decisions.

## Conclusions

This study is the first to extensively evaluate in silico tool performance in multiple solid tumors with actionable genetic targets. We show MutationTaster2021 as the highest-performing tool, outperforming meta-predictor tools that have demonstrated superior performance in previous studies. Our results support that in silico classification tools may offer valuable insights into confirming or ruling out pathogenicity of VUS. However, they should be approached cautiously, acknowledging the substantial variability in pathogenicity predictions.

## Data Availability

All data produced in the present study are available upon reasonable request to the authors

## Acknowledgements

Not applicable.

