## Supplementary Material for "Evaluation of *in silico tools* for variant classification in missense variants of solid cancer with actionable genetic targets"

**Table 1. Overall In Silico Tool Performance and In Silico Tool Performance Across Cancer Types**

| **Algorithm** | **Total variants (n)** | **Accuracy** | **Sensitivity** | **Specificity** | **PPV** | **NPV** | **MCC** |
| --- | --- | --- | --- | --- | --- | --- | --- |
| **Overall Performance (n = 1161)** | | | | | | | |
| Align-GVGD | 1140/1161 | 0.555 | 0.843 | 0.242 | 0.548 | 0.587 | 0.107 |
| CADD | 1159/1161 | 0.666 | 0.983 | 0.319 | 0.612 | 0.947 | 0.411 |
| FATHMM | 1146/1161 | 0.655 | 0.775 | 0.523 | 0.642 | 0.679 | 0.309 |
| MutationTaster2021 | 1149/1161 | 0.829 | 0.927 | 0.721 | 0.784 | 0.900 | 0.666 |
| PolyPhen-2 (HumDiv) | 1149/1161 | 0.646 | 0.834 | 0.439 | 0.621 | 0.706 | 0.293 |
| PolyPhen-2 (HumVar) | 1149/1161 | 0.653 | 0.738 | 0.559 | 0.648 | 0.659 | 0.302 |
| REVEL | 1160/1161 | 0.702 | 0.851 | 0.539 | 0.668 | 0.769 | 0.413 |
| **Breast Cancer (n = 823)** | | | | | | | |
| Align-GVGD | 810/823 | 0.500 | 0.855 | 0.237 | 0.454 | 0.688 | 0.114 |
| CADD | 821/823 | 0.626 | 0.986 | 0.362 | 0.523 | 0.972 | 0.418 |
| FATHMM | 809/823 | 0.618 | 0.649 | 0.595 | 0.544 | 0.695 | 0.241 |
| MutationTaster2021 | 814/823 | 0.857 | 0.930 | 0.804 | 0.777 | 0.940 | 0.726 |
| PolyPhen-2 (HumDiv) | 811/823 | 0.614 | 0.832 | 0.453 | 0.530 | 0.784 | 0.299 |
| PolyPhen-2 (HumVar) | 811/823 | 0.626 | 0.678 | 0.588 | 0.549 | 0.712 | 0.264 |
| REVEL | 822/823 | 0.674 | 0.810 | 0.574 | 0.583 | 0.805 | 0.386 |
| **Ovarian Cancer** | | | | | | | |
| Align-GVGD | 693/706 | 0.466 | 0.867 | 0.233 | 0.397 | 0.750 | 0.121 |
| CADD | 704/706 | 0.601 | 0.988 | 0.377 | 0.478 | 0.982 | 0.410 |
| FATHMM | 692/706 | 0.585 | 0.537 | 0.613 | 0.448 | 0.694 | 0.146 |
| MutationTaster2021 | 699/706 | 0.858 | 0.907 | 0.830 | 0.756 | 0.939 | 0.716 |
| PolyPhen-2 (HumDiv) | 694/706 | 0.598 | 0.835 | 0.460 | 0.473 | 0.828 | 0.298 |
| PolyPhen-2 (HumVar) | 694/706 | 0.622 | 0.671 | 0.595 | 0.490 | 0.757 | 0.256 |
| REVEL | 704/706 | 0.689 | 0.899 | 0.567 | 0.546 | 0.907 | 0.460 |
| **Pancreatic Cancer** | | | | | | | |
| Align-GVGD | 456/457 | 0.522 | 0.872 | 0.220 | 0.491 | 0.667 | 0.121 |
| CADD | 457/457 | 0.582 | 0.976 | 0.244 | 0.526 | 0.923 | 0.314 |
| FATHMM | 455/457 | 0.571 | 0.550 | 0.590 | 0.537 | 0.602 | 0.140 |
| MutationTaster2021 | 454/457 | 0.815 | 0.895 | 0.745 | 0.752 | 0.892 | 0.643 |
| PolyPhen-2 (HumDiv) | 457/457 | 0.586 | 0.853 | 0.358 | 0.533 | 0.739 | 0.239 |
| PolyPhen-2 (HumVar) | 457/457 | 0.580 | 0.692 | 0.484 | 0.535 | 0.647 | 0.179 |
| REVEL | 457/457 | 0.643 | 0.890 | 0.431 | 0.573 | 0.822 | 0.357 |
| **Prostate Cancer** | | | | | | | |
| Align-GVGD | 453/454 | 0.519 | 0.871 | 0.214 | 0.489 | 0.658 | 0.125 |
| CADD | 454/454 | 0.581 | 0.976 | 0.242 | 0.526 | 0.922 | 0.312 |
| FATHMM | 452/454 | 0.573 | 0.552 | 0.591 | 0.540 | 0.603 | 0.143 |
| MutationTaster2021 | 451/454 | 0.814 | 0.895 | 0.744 | 0.751 | 0.891 | 0.640 |
| PolyPhen-2 (HumDiv) | 454/454 | 0.588 | 0.852 | 0.361 | 0.534 | 0.739 | 0.242 |
| PolyPhen-2 (HumVar) | 454/454 | 0.581 | 0.690 | 0.488 | 0.537 | 0.647 | 0.181 |
| REVEL | 454/454 | 0.645 | 0.890 | 0.434 | 0.575 | 0.822 | 0.359 |
| **Colorectal Cancer** | | | | | | | |
| Align-GVGD | 127/127 | 0.709 | 0.828 | 0.286 | 0.804 | 0.320 | 0.119 |
| CADD | 127/127 | 0.780 | 1.00 | 0.00 | 0.780 | Undefined | 0.00 |
| FATHMM | 126/127 | 0.778 | 1.00 | 0.00 | 0.778 | Undefined | 0.00 |
| MutationTaster2021 | 125/127 | 0.808 | 0.990 | 0.148 | 0.808 | 0.800 | 0.290 |
| PolyPhen-2 (HumDiv) | 127/127 | 0.756 | 0.808 | 0.571 | 0.870 | 0.457 | 0.352 |
| PolyPhen-2 (HumVar) | 127/127 | 0.772 | 0.828 | 0.571 | 0.872 | 0.485 | 0.378 |
| REVEL | 127/127 | 0.843 | 0.970 | 0.393 | 0.850 | 0.786 | 0.480 |
| **Bladder Cancer** | | | | | | | |
| Align-GVGD | 100/100 | 0.610 | 0.810 | 0.270 | 0.654 | 0.455 | 0.093 |
| CADD | 100/100 | 0.650 | 0.984 | 0.081 | 0.646 | 0.750 | 0.161 |
| FATHMM | 100/100 | 0.630 | 0.889 | 0.189 | 0.651 | 0.500 | 0.109 |
| MutationTaster2021 | 100/100 | 0.650 | 0.889 | 0.243 | 0.667 | 0.563 | 0.174 |
| PolyPhen-2 (HumDiv) | 100/100 | 0.600 | 0.794 | 0.270 | 0.649 | 0.435 | 0.073 |
| PolyPhen-2 (HumVar) | 100/100 | 0.590 | 0.746 | 0.324 | 0.653 | 0.429 | 0.076 |
| REVEL | 100/100 | 0.650 | 0.857 | 0.297 | 0.675 | 0.550 | 0.186 |
| **Biliary Cancer** | | | | | | | |
| Align-GVGD | 83/91 | 0.566 | 0.776 | 0.265 | 0.603 | 0.450 | 0.046 |
| CADD | 91/91 | 0.637 | 0.982 | 0.059 | 0.636 | 0.667 | 0.112 |
| FATHMM | 91/91 | 0.626 | 0.877 | 0.206 | 0.649 | 0.500 | 0.111 |
| MutationTaster2021 | 91/91 | 0.626 | 0.877 | 0.206 | 0.649 | 0.500 | 0.111 |
| PolyPhen-2 (HumDiv) | 91/91 | 0.582 | 0.789 | 0.235 | 0.634 | 0.400 | 0.029 |
| PolyPhen-2 (HumVar) | 91/91 | 0.571 | 0.754 | 0.265 | 0.632 | 0.391 | 0.021 |
| REVEL | 91/91 | 0.615 | 0.842 | 0.235 | 0.649 | 0.471 | 0.096 |
| **Melanoma** | | | | | | | |
| Align-GVGD | 87/87 | 0.724 | 0.866 | 0.250 | 0.795 | 0.357 | 0.132 |
| CADD | 87/87 | 0.770 | 1.00 | 0.00 | 0.770 | Undefined | 0.00 |
| FATHMM | 87/87 | 0.770 | 1.00 | 0.00 | 0.770 | Undefined | 0.00 |
| MutationTaster2021 | 85/87 | 0.824 | 1.00 | 0.211 | 0.815 | 1.00 | 0.414 |
| PolyPhen-2 (HumDiv) | 87/87 | 0.736 | 0.791 | 0.550 | 0.855 | 0.440 | 0.317 |
| PolyPhen-2 (HumVar) | 87/87 | 0.759 | 0.821 | 0.550 | 0.859 | 0.478 | 0.354 |
| REVEL | 87/87 | 0.816 | 0.955 | 0.350 | 0.831 | 0.700 | 0.403 |
| **Thyroid Cancer** | | | | | | | |
| Align-GVGD | 55/55 | 0.691 | 0.814 | 0.250 | 0.795 | 0.273 | 0.066 |
| CADD | 55/55 | 0.782 | 0.953 | 0.167 | 0.804 | 0.500 | 0.191 |
| FATHMM | 55/55 | 0.782 | 1.00 | 0.00 | 0.782 | Undefined | 0.00 |
| MutationTaster2021 | 55/55 | 0.800 | 0.930 | 0.333 | 0.833 | 0.571 | 0.327 |
| PolyPhen-2 (HumDiv) | 55/55 | 0.745 | 0.907 | 0.167 | 0.796 | 0.333 | 0.098 |
| PolyPhen-2 (HumVar) | 55/55 | 0.764 | 0.907 | 0.25 | 0.813 | 0.429 | 0.195 |
| REVEL | 55/55 | 0.800 | 0.930 | 0.333 | 0.833 | 0.571 | 0.327 |

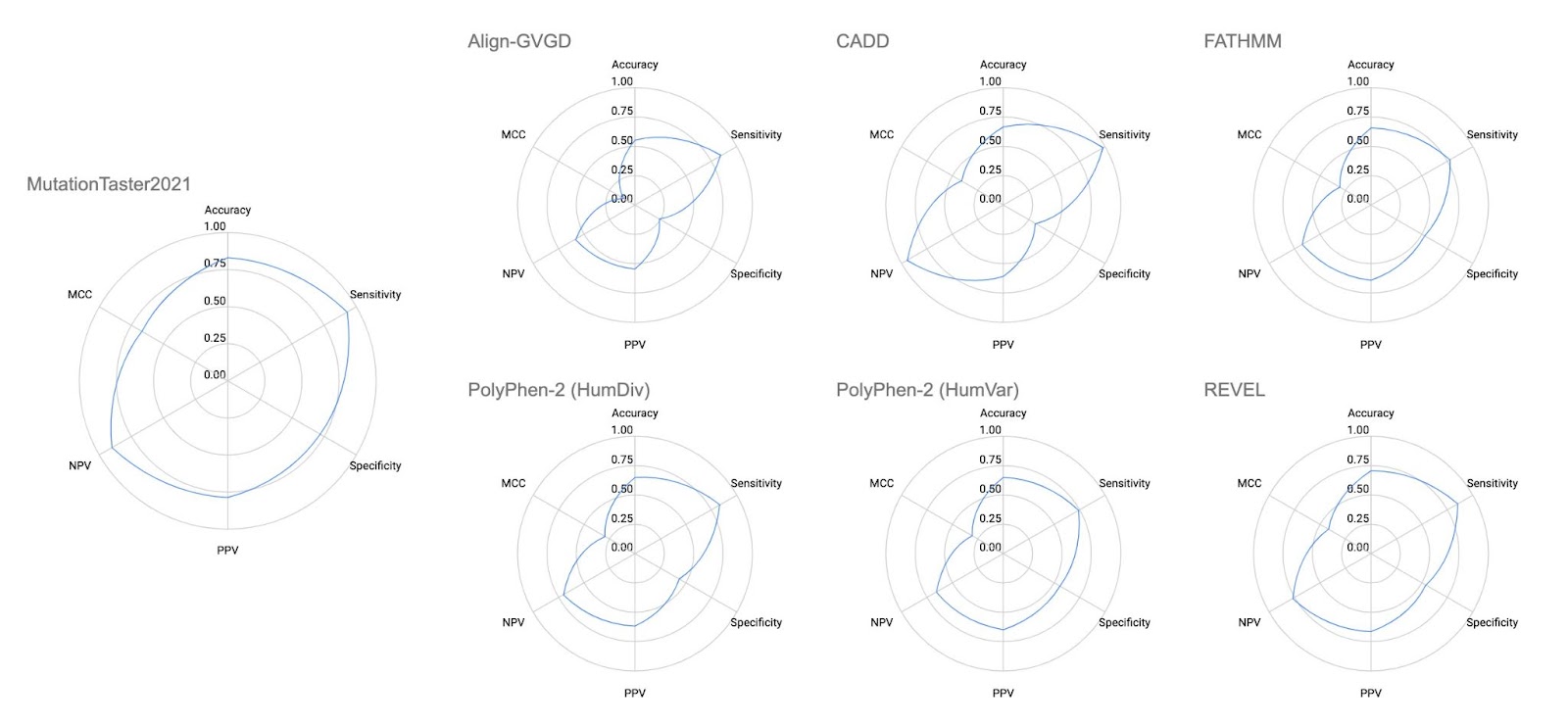

C

G

F

E

D

B

A

**Figure 1.** Radar charts demonstrating overall performance according to tool. (A) MutationTaster2021 exhibited the highest overall performance, with the highest scores in accuracy, specificity, PPV, and MCC. (B) Align-GVGD exhibited the lowest overall performance, with the lowest scores in accuracy, specificity, PPV, NPV, and MCC. (C) PolyPhen-2 (HumDiv), (D) CADD, (E) PolyPhen-2 (HumVar), (F) FATHMM, and (G) REVEL were other tools examined, but notable performance was not observed.
